# Epidemiology and predictors of repeat positive chlamydia tests: The Brant County cohort, Ontario, Canada

**DOI:** 10.1101/19007278

**Authors:** Jenny Pereira Santos, Alexey Babayan, Miao Jing Huang, Ann Jolly

## Abstract

**Objectives:** Repeat positive tests for chlamydia (CR) may help explain current high rates of chlamydia despite years of screening, partner notification and treatment to reduce sequelae. We wanted to determine the numbers of CRs over time as a proportion of all chlamydia cases, and define the differences in demographic, clinical, behavioural, and public health management indicators, between individuals who have experienced a CR and individuals who experienced a single infection in Brant County, Ontario.

**Methods:** A retrospective cohort was developed using notifiable disease data extracted from the integrated public health system. Cases were laboratory confirmed chlamydia and gonorrhea infections in Brant County between January 1^st^, 2006 and December 31^st^, 2015. During the study period, 3,499 chlamydia cases and 475 gonorrhea cases were diagnosed. The total number of individuals with chlamydia in that period was 3,060, including 157 coinfections with gonorrhea. Differences between those with reinfection and those with single infection were evaluated using univariate and multivariate (Cox proportional hazards model) methods.

**Results:** Four hundred and ninety-nine (16.30%) individuals experienced CR 28 days from initial infection; of which 328 (65.73%) occurred within 2 years and 211 (42.28%) within 1 year. The median time to CR was 276 days, consistent with existing Canadian literature. Independent risk factors for CR included being male, 25 years old or younger, and not receiving recommended treatment for initial and/or subsequent infection.

**Conclusions:** These findings suggest that inadequate treatment play a significant role in CR, while accounting for young age and male gender, likely due to untreated sex partners.

**Key Messages:**

- Sixteen percent of people experienced a second positive chlamydia test more than 28 days after their initial positive test in a cohort of 3,499 patients
- Those who had a second positive test were more likely to be male, younger than 25 and had not received recommended antimicrobials
- Confirmation of any kind of partner notification was missing in 88% of records

## INTRODUCTION

Screening for chlamydia treatment and partner notification were initially introduced in Canada in the late 1980’s-in the absence of randomized clinical trials (RCTs)-in order to reduce pelvic inflammatory disease (PID), ectopic pregnancy and infertility.^1^ When chlamydia was made notifiable, screening women younger than 25 was recommended with other groups at risk and treatment and sex partner management guidelines were included. Initially, chlamydia decreased to a low of 129/100,000 in 1998, after which it more than doubled by 2016 (334/100,000).^2^ The rise has been attributed to factors such as more risky sex behaviours,^3,4^ improvements in test sensitivity, increased screening of men,^5^ increased testing and rescreening of women ^4,6^ and arrested immunity.^3,4,7^ It is well established that the epidemiology of sexually transmitted infections (STIs) changes under the pressure of control programs. Interventions that were cost efficient and effective in the initial phases may need to be reviewed in favour of more strategic, targeted approaches focusing on the core group of people with STI who, together with their partners contribute disproportionately to STI transmission.^8,9^

A recent Cochrane review of RCT of chlamydia screening for reduction of sequelae showed conflicting evidence from high-quality studies. Two studies showed no reduction in PID rates following screening, while another two were able to show higher rates of PID in screened women. ^10^ A Canadian cohort study confirmed an increased dose-response risk of PID following one, two, three or more positive tests for chlamydia over those who tested negative.^11^ This may reflect the fact that the risk of PID also increases in women proportionate to duration of infection.^12^ Although men and women are just as likely to contract the infection, incidence rates in females appear higher than those in males, due in part to a larger portion of females being tested. However, males have higher positivity rates than females; suggesting that many are under diagnosed and constitute a “hidden reservoir” of infection.^5,13^ Accordingly, investigations into the increases in bacterial STIs was ranked as the leading research priority in bacterial sexually transmitted infections (STIs) in Canada. ^14^

Brant County is located in central Ontario and had a population of approximately 125,099 in 2006, which increased to approximately 129,288 by 2011. The incidence rate for chlamydia in the area was 150/100,000 in 2006, which more than doubled by 2011 (395/100,000),^15^ compared to only a 54% increase in incidence in Ontario and a 38% increase within Canada.^16^ The majority of chlamydia infections reported to the Brant County Health Unit (BCHU) were in adults between 20 and 24-years-old, followed by females between 15 and 19-years-old and males between 25 and 29-years-old,^15^ similar to the epidemiology in Ontario, Canada and elsewhere.^4,15,17–19^

The Public Health Standards in Ontario for bacterial STI control focus mainly on screening, re-testing cases, and partner notification/contact tracing.^20^ When people are diagnosed, they are reported to the Ministry of Health for registration, treatment, and follow-up after 6 months.^20^ Here, we assess the proportion of repeat positive tests, comprising genuine reinfections and continuing infections (CR) of all chlamydia positive tests in Brant County; estimated to be between 6 – 10% in other Canadian cities.^5,21^ We hypothesize that individuals who experience a CR within two years after initial infection differ significantly from individuals with single infections in terms of demographics, risk factors, reasons for testing, treatment, and contact tracing indicators.

## METHODS

### Study Design and Population

A population-based retrospective cohort was constructed using data extracted from the integrated public health information system (iPHIS), for the period between January 1^st^, 2006 and December 31^st^, 2015. iPHIS is used by public health units to report cases of notifiable diseases to Public Health Ontario, in accordance to the Health Protection and Promotion Act.^22^ An iPHIS record is generated on receipt of; a computerized positive laboratory reports of notifiable sexually transmitted infections; a physician report, or a notification from outside of Brant County of a positive case.^22^ An assessment of positive *N. meningitidis* Serogroup C,^23^ and positive pertussis PCR^24^ laboratory records revealed that 91% and 84%, respectively were present in iPHIS, though not all laboratory positive cases met the case surveillance definition of a pertussis case. It is the most complete single source of validated, infectious disease reports and has been used as a gold standard in studies of other notifiable infections.^23,25^ All individuals with a laboratory confirmed *C. trachomatis* infection in Brant County from 2006 onwards were followed until they either experienced a CR or until they reached the end of the study period. Extracted information included age at time of positive test, residential address, risk behaviours, reasons for testing, clinical information (such as diagnosis date, record of past infections, treatment information) and whether or not contact tracing had taken place. Data on laboratory confirmed *Neisseria gonorrhoeae* infections were also included in order to study and control for coinfection. All other bacterial STI diagnoses were excluded. The Ottawa Health Science Network Research Ethics Board approved this study (certificate number 2015067901H.)

### Measures

A CR was defined as a second diagnosis occurring 28-730 (2 years) days after the initial infection. We were unable to differentiate reinfections from continuing infections due to lack of complete treatment and negative test data; only the first CR was included in analysis. Individuals without a second positive test, or who moved out of Brant, were censored at the end of the study period. Age, sex, evidence of partner notification, risk factors, reason for testing (including routine screening and symptoms), and postal code (used to determine whether cases were located within the previously identified core group in downtown Brantford) were included in the final model. The age of cases was analyzed as both a continuous and a categorical variable. Risk factors were grouped into three categories: 1) high risk-behavioral (anonymous sex, more than one sex contact in the last 6 months, new contact in past 2 months, no condom used, alcohol or injection/inhalation drug use, sex trade worker or homeless), 2) high risk-medical (repeat STI, pregnant or HIV), and 3) low risk/other factors (factors such as bath house use, travel, having been in a correctional facility, and condom breakage). Most information on risk factors and reasons for testing were missing (n=3,060), therefore we also compared complete and incomplete records to investigate if data were missing at random.

### Analysis

The Mantel-Haenszel *X*^2^ test was used for bivariate analysis to determine the relationship between categorical variables. The Cox proportional-hazards model was used to determine the influence of multiple covariates on time to CR (measured in number of days). Covariates included: age, sex, treatment received, partner notification, reason for testing, and coinfection. The proportional-hazards assumptions that survival curves have hazard functions that are constant over time and that censoring of an individual is unrelated to the probability of a CR occurring, were not violated. All analyses were conducted using SAS version 9.3 (SAS Institute Inc., Cary, NC).

## RESULTS

### Descriptive Statistics

There were 7,654 index cases of chlamydia and gonorrhea identified in Brant County within the study period. Of these, 3,499 chlamydia cases (representing 3,000 unique individuals) remained after removing duplicate records and those with gonorrhea only. Sixteen percent (499) of the 3,000 individuals experienced a CR after 28 days after their initial infection and 66% percent (328) occurred within two years. Characteristics of individuals with a CR that occurred within one year were not significantly different from those who experienced a CR within two years, (not shown). The CR count rose sharply between 2009 and 2010 from 23 (3% of chlamydia cases), to 50 in 2012 (11% of cases), consistent with a testing campaign (Figure 1). Even though it began to fall following 2012, it remained high in comparison to other areas in Canada^26^ until most recently in 2015. The median time to CR was 276 days, (about 9 months).

**Figure 1.**
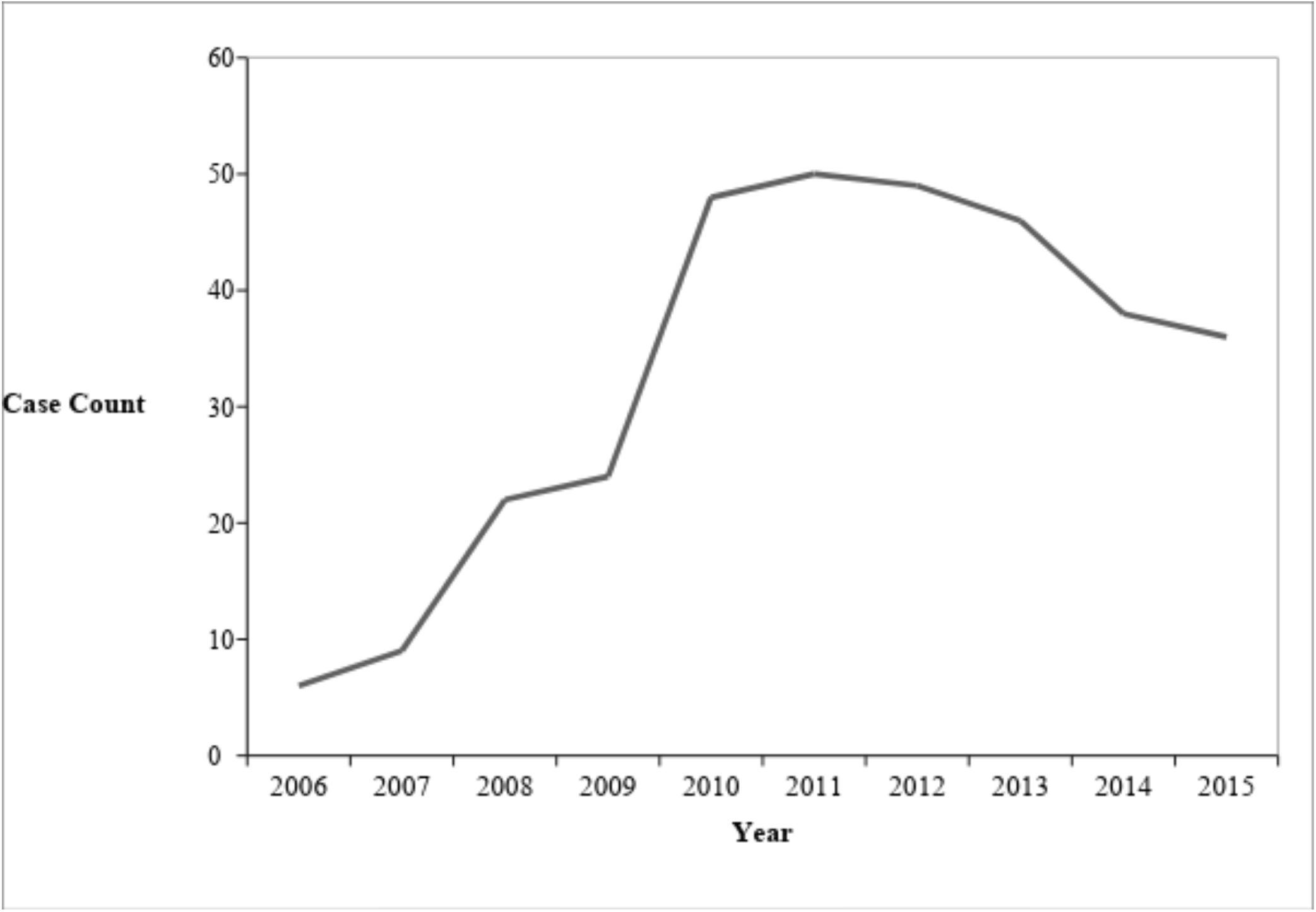
Case Count of CR within two years of a first chlamydia infection in Brant County, Ontario, 2006-2015 (n= 328)

Although most CR cases occurred in females (75%), the rate of CR was nearly the same between sexes; 12% of females and 8% of males experienced a CR within two years. Of the CR cases, 60 (12.02%) were also co-infected with gonorrhea. When cases with gonorrhea were included to determine the extent of coinfection, the final sample size was 3,060 cases, which included 167 cases of coinfection, representing 2,829 unique individuals.

The median age among all cases was 23 (median age for coinfected only and CR only were 21.50 and 21.15, respectively). As expected, individuals within the age range 18-25 accounted for the most initial and CR cases (see Table 1). The majority of cases were females (75%) and were reported as having been prescribed the recommended treatment of 1gm azithromycin and/or 100mg doxycycline (92%); 177 (6.3%) had no treatment record.

**Table 1.**
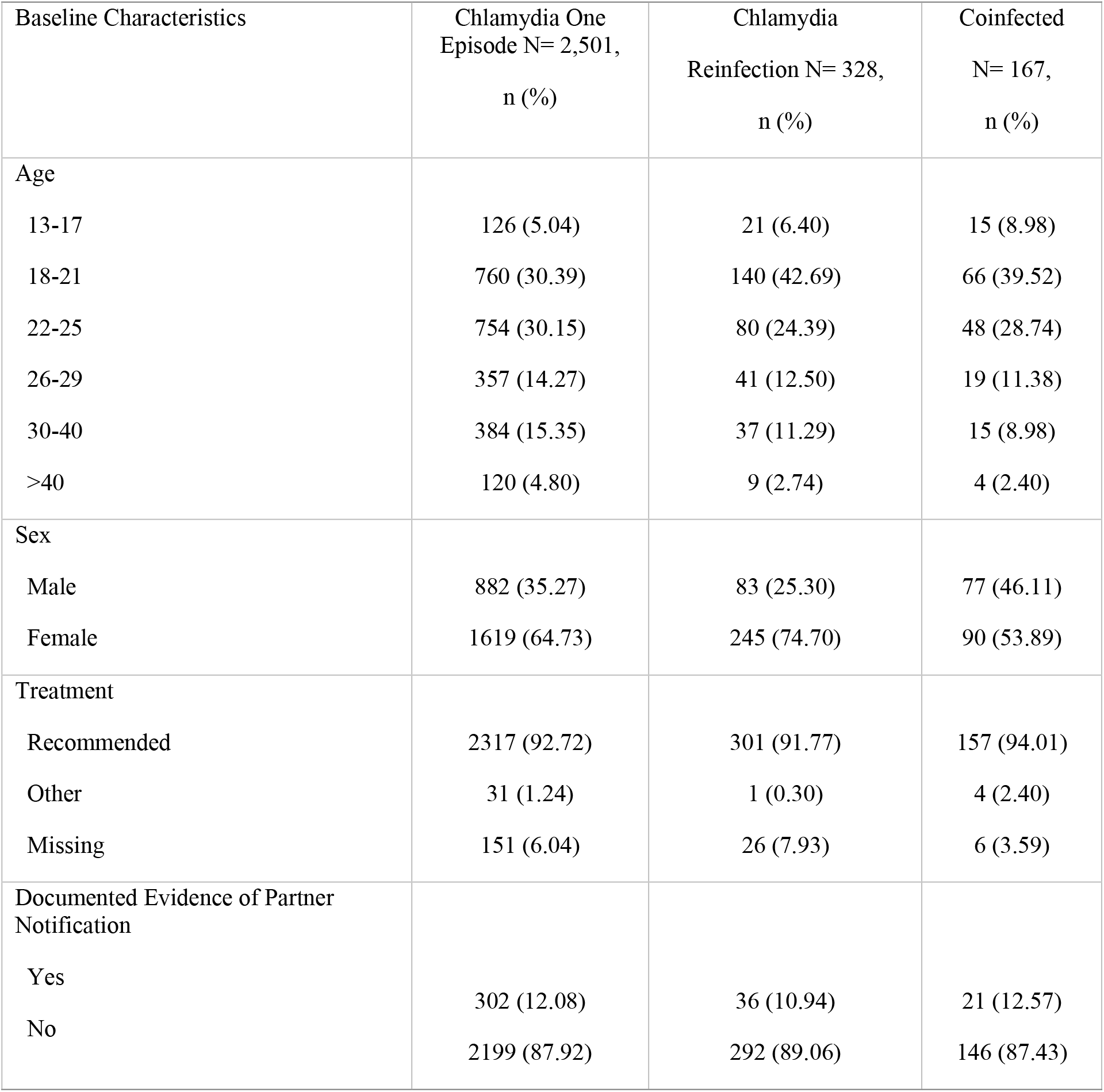
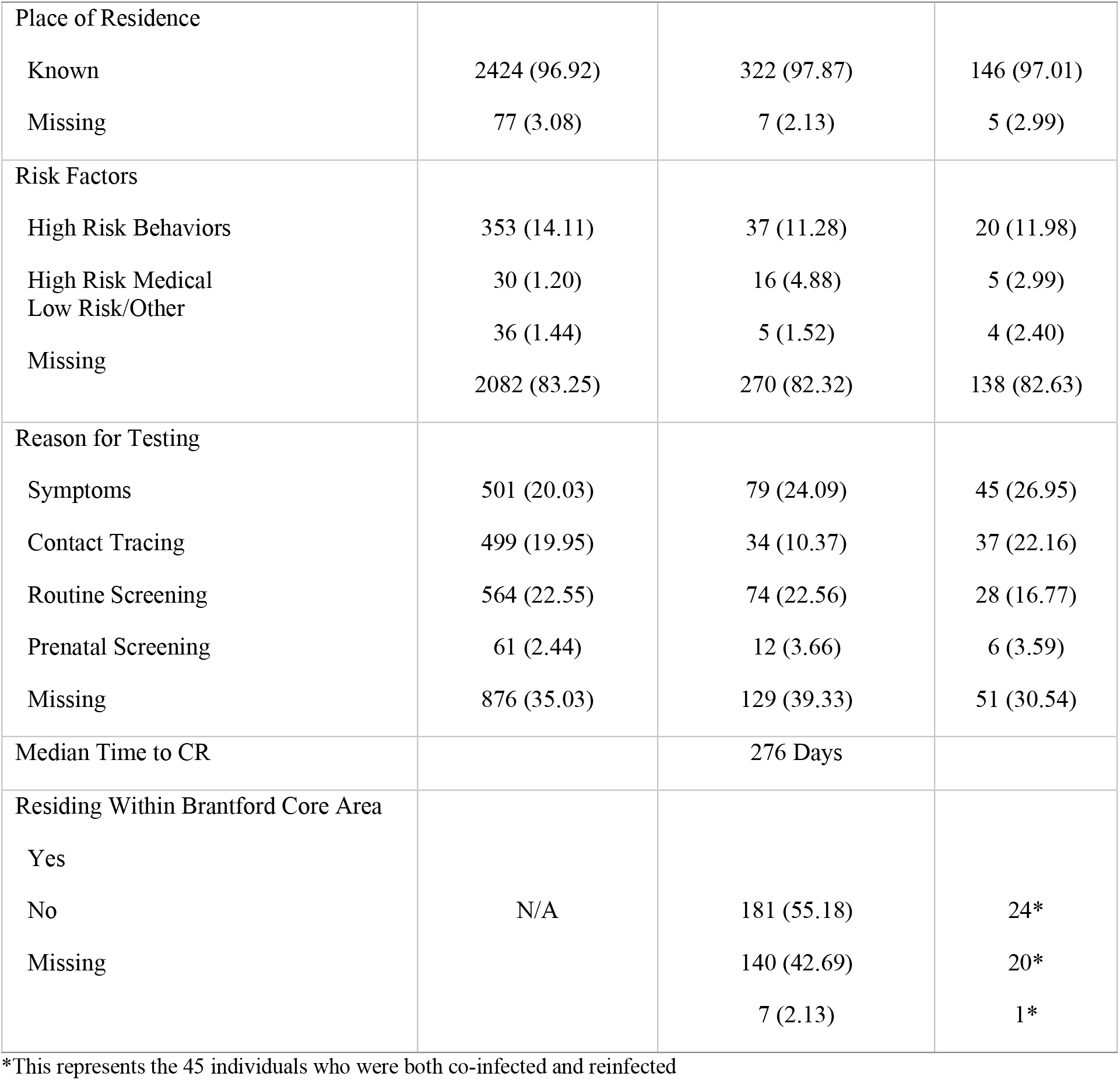
Baseline characteristics of individuals with chlamydia reinfections and coinfection in comparison to those with single infections, 2006-2015, Brant County, Ontario (n=2,829)

Partner notification was documented for only 364 (12.13%) individuals overall (and 36 or 10.94% of CR cases). Of the CR cases, 60 (12.02%) were also co-infected with gonorrhea and of those, partner notification was completed for 45 individuals (13.72%). *X*^2^ tests indicated that coinfection was significantly associated with CR (p= <.0001) but partner notification was not (p= 0.4420). Of CR cases, 7 (2.13%) had missing residential information and 2 (0.613%) had a residential address in areas outside of Brant County. The latter were removed from the final model, without a significant change in the results.

Overall, routine screening (22.27%) was the most common reason for testing, followed by having symptoms (20.87%). A *X*^2^ test indicated that providing a “reason for testing” was significantly associated with CR (p= 0.0027). The most common risk factor cited was “no condom use”, but no single risk factor was associated with CR in the bivariate analysis (p= 0.35). As a large proportion of the study population (83.13%) did not provide risk factor information, this variable was excluded from the final model. There were no evident demographic differences between cases with missing data and those without (Table 2). However, CR occurred sooner in those missing reasons for being tested.

**Table 2.** Characteristics of chlamydia infections by risk factor information (available vs. unavailable), 2006-2015, Brant County, Ontario (n=3,060)

| Baseline Characteristics | Risk Factor Information<br>Available,<br>n= 525<br>n (%) | Risk Factor Information<br>Unavailable,<br>n= 2535<br>n (%) |
| --- | --- | --- |
| Age |  |  |
| 13-17 | 29 (5.52) | 123 (4.85) |
| 18-21 | 158 (30.10) | 801 (31.60) |
| 22-25 | 160 (30.48) | 760 (29.98) |
| 26-29 | 71 (13.52) | 372 (14.67) |
| 30-40 | 83 (15.81) | 368 (14.52) |
| >40 | 24 (4.57) | 111 (4.38) |
| Sex |  |  |
| Male | 165 (31.43) | 861 (33.96) |
| Female | 360 (68.57) | 1674 (66.04) |
| Treatment* |  |  |
| Recommended | **508 (96.76)** | **2328 (91.83)** |
| Other | **9 (1.71)** | **25 (0.99)** |
| Missing | **8 (1.52)** | **182 (7.18)** |
| Partner Notification* |  |  |
| Yes | **317 (60.38)** | **54 (2.13)** |
| No | **208 (39.62)** | **2481 (97.87)** |
| Place of Residence |  |  |
| Known | 520 (99.05) | 2452 (96.73) |
| Missing | 5 (0.95) | 83 (3.27) |
| Reason for Testing* |  |  |
| Symptoms | **162 (30.86)** | **472 (18.62)** |
| Contact Tracing | **105 (20.00)** | **468 (18.46)** |
| Routine Screening | **137 (26.10)** | **555 (21.89)** |
| Prenatal Screening | **36 (6.86)** | **48 (1.89)** |
| Missing | **85 (16.19)** | **992 (39.13)** |
| Reinfection |  |  |
| Yes | 106 (20.19) | 453 (17.87) |
| No | 419 (79.81) | 2082 (82.13) |
| Mean Time to CR* | **165 Days** | **108 Days** |
| Coinfection |  |  |
| Yes | 9 (1.71) | 51 (2.01) |
| No | 516 (98.29) | 2484 (97.99) |

\* Indicates statistically significant differences

### Survival Analysis

Table 3 shows the multivariate Cox regression model. Independent risk factors for CR included being 25 years old or younger (HR= 3.19, 2.77, 1.61) and not receiving recommended antimicrobial treatment (HR= 1.44). With every year increase in age (analyzed as a continuous variable) an individual had a 6.1% decrease in risk of CR (p= <.0001). However, being female was protective (HR= 0.79), meaning males were more likely to present with a CR than were females (HR=1.26).

**Table 3.**
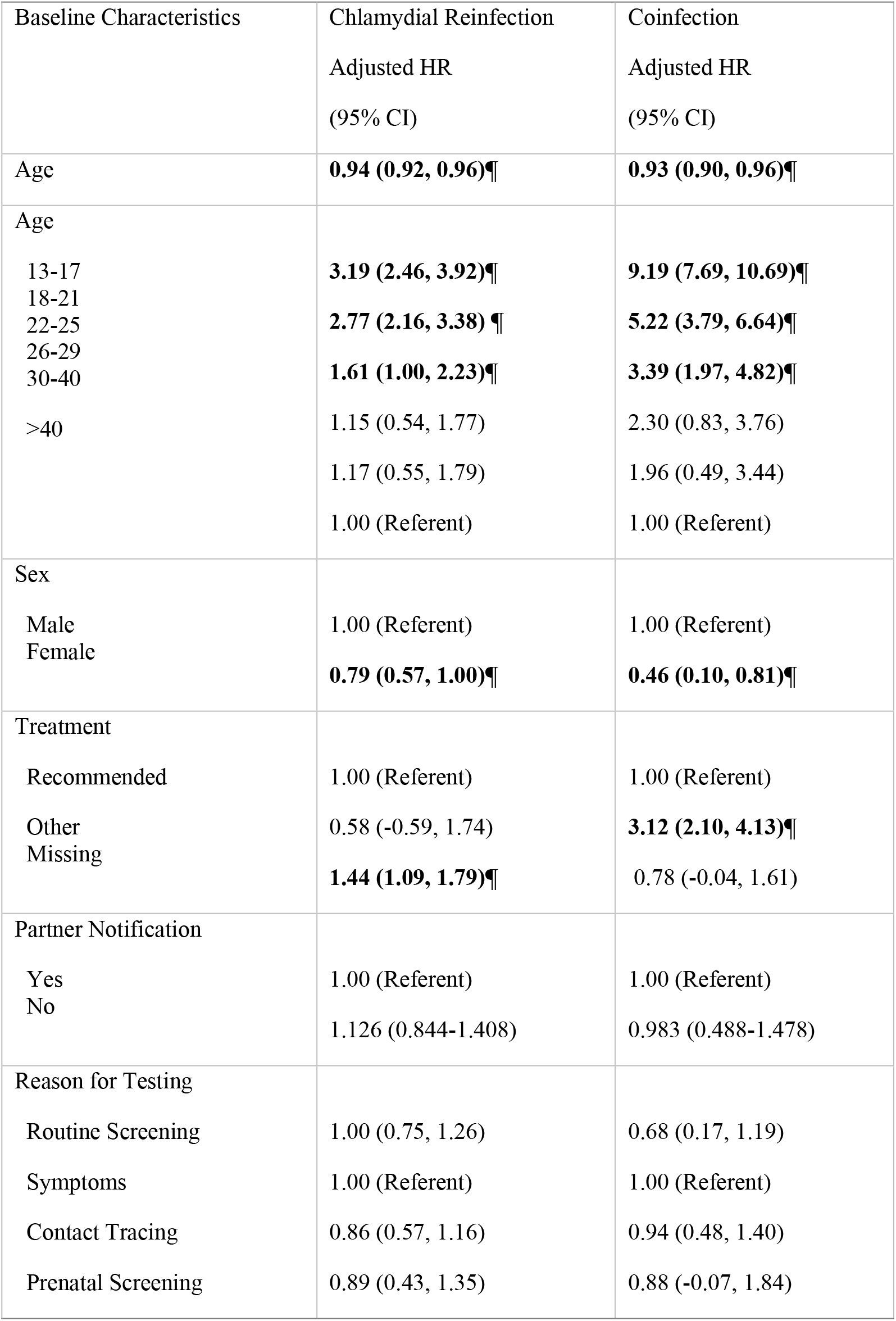
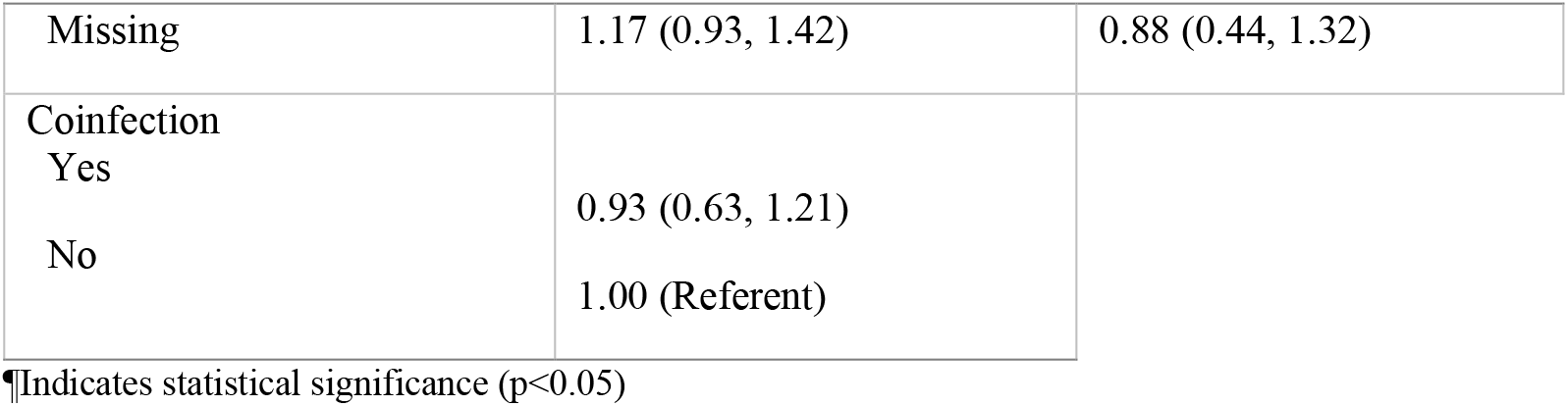
Multivariate Cox regression analysis of time to CR, by baseline characteristics, two-year follow-up, 2006-2015, Brant County, Ontario (N= 3,060)

Although coinfection was not a statistically significant risk factor for CR (p= 0.52), the final survival analysis model was run a second time with coinfection as the outcome of interest, rather than a risk factor, as they were still found to be associated with one another (P=<0.01). The statistically significant risk factors for coinfection were similar to those for CR, being 25 or younger and not receiving the recommended antimicrobial treatment. Similar to the CR model, being female had a protective effect on coinfection.

## DISCUSSION

Over time, the rate of CR rose by a factor of 10 from approximately 5 women in 2006 to 50 in 2011, when a chlamydia test campaign was launched, (personal communication Shawna Wilson, May 30, data not shown) after which it dropped to about 40 cases a year. Numbers of tests followed the same trend, with an initial rise of 30% from 2005 - 2011 after which they dropped back to previous levels. The percent of positive tests doubled over the period, from 3.4 – 7%, with the highest proportions in 2011, indicating a real rise in infection rather than an artifact of testing. This indicates also that CR is at least partly responsible for the rise in chlamydia rates in Brant County. The rise coincides with the Ontario recommendations in 2008 to rescreen women 6 months after a positive test, and those with high risk behaviours every three months. Although routine screening (i.e. testing for infection in a person without signs or symptoms) was the most common reason given for testing in this population, it was not distinguished from diagnostic testing within iPHIS, so it is possible that people who had symptoms were incorrectly reported as “screened.”^27^ This may be resolved by providing separate fields for responses to questions on reason for testing, so that more than one reason can be entered, allowing public health staff to differentiate screening from diagnostic testing.^28^

We found that 328 individuals experienced a CR within two years of initial infection (median= 9 months) between 2006 - 2015, which represents a 10.9% rate of CR. This is consistent with both Canadian^5,26^ and American literature.^29^ Similar to other studies, time to CR was associated with young age.^5,26^ Males were more likely to be to be diagnosed with CR due to the fact that they are more likely to experience symptoms and then be tested, and they have lower partner testing and treatment rates than women, making re-exposure after their treatment more likely, as in a previous study.^13^ Unlike previous studies, neither partner notification nor coinfection was significantly associated with time to CR, although in a cross sectional study chlamydia repeaters had higher median numbers of reported partners than those with only one chlamydia diagnosis.^13^ This is likely due to the fact that overall adherence to partner notification protocols was poor in the current data, with only 10% of cases reported to undergo the process. Also, STI treatment guidelines recommend automatic treatment for chlamydia in those positive or suspected to have gonorrhea. As gonorrhea is more frequently symptomatic, this prompts people to present more frequently, at which time they treated for both chlamydia and gonorrhea and thus making CR less likely. Most important, individuals whose records were missing treatment data were more 1.44 times more likely to experience a CR, identical to the OR of a previous cross-sectional comparison.^13^ This either indicates that treatment had not been prescribed, or that it was received in a physician’s office, and not at the STI clinic, which increased the likelihood that the treatment prescribed may not have been in accordance with the current recommendations, borne out by a study in which researchers found only 30-60% compliance by physicians with Canadian STI treatment recommendations.^30^

The lack of documentation on partner notification undertaken by health care providers or patients themselves, prompts questions about the relationship between screening, rescreening and/or retesting women who may be re-exposed to an untreated partner. It is possible, for example, that resources spent on screening and rescreening patients may detract from those required to provide recommended thorough patient and partner education and care. We did construct sexual networks from reported partner notification data, which were small and sparse, with only 364 (12%) cases who nominated partners, which resulted in many unattached individuals and 44 components ranging in size from 2-12 people and only 4 exceeding 4 people. That only 10% or less of infected patients’ partners received follow-up represents a pervasive erosion of partner services and reporting of follow-up over time.^31^

As positive chlamydia tests are continuing to increase, reinfections form at least 10% of the rise. Because recent studies show that (1) 20% of infections in women resolve on their own; (2) women with self-resolved infection are four times less likely to be reinfected;^32^ (3) the percentage of positive tests of all those tested is decreasing, altering the cost for each case found,^6^ and (4) Cochrane review of high quality studies produced no evidence that screening prevents sequelae,^33^ we believe that detailed monitoring and evaluation of STI care is vital. It is also possible that due to changes in management of cases, the proportion chlamydia repeaters who constitute the chlamydia core group,^13^ have increased relative to the number of those testing positive. This is likely due to longer durations of infectiousness as below, resulting in higher rates overall. The importance of core groups in evolving epidemics of STI strongly reinforces the need to collect essential data for targeted prevention.

We specifically analyzed the patterns of risk behaviors and reasons for testing and found a substantial amount missing. Individuals missing this information were; more likely to be inadequately treated, less likely to have partners notified, and less likely to have reasons for testing recorded. Most important, individuals who were missing this information had a shorter mean time to CR compared to those who had complete information (108 and 165 days, respectively), as above, suggesting that the quality of care, like the documentation, may be suboptimal.

While patients may be reluctant to provide information on partners, risk interactions, and reasons for testing, to healthcare professionals, the providers working in sexual health may be inadequately resourced, trained and/or supported to conduct those discussions well, and/or are unaware of the importance of this client information for sound patient management and program evaluation. However, information is available for data elements, and system architecture required for case management, surveillance, notifiable disease purposes evaluation of STI control has been specified, and will facilitate the refinement of interventions for people at high risk.^28^

Following this research, walk-in services were provided in the area of the county that was identified as being at the highest risk (the core group) in downtown Brantford, to facilitate screening, diagnosis and treatment, thus reducing infectious periods. More complete information on risk factors and partner notification is being collected in Brant County which will indicate whether options such as adding a web-based partner notification system and/or expedited partner delivered therapy (EPT) may be effective in this population. Methods that strengthen partner notification procedures have been found to contribute as much to chlamydia control as methods used to strengthen screening procedures,^34,35^as does EPT.^36–38^ However, this would be only beneficial if CR cases are true reinfections rather than treatment failure or inadequacy. In addition, we will also evaluate screening coverage, which facilitate better understanding of the proportion and nature of individuals who are undergoing screening, their positivity rates, and the risk of CR.

In conclusion, we focused on the epidemiology of CR so as to better define a possible cause of some of current increases in chlamydia. The low number of reports of partner follow-up, repeated exposures to untreated partners, subsequent positive tests, missing treatment data and reasons for infection reinforced the fact that complete, accurate data for STI surveillance and management and active program evaluation are essential when the effectiveness of screening for chlamydia is in question.

## Data Availability

The data used in this study are not available for public use, as they contain identifying information including age, sex and dissemination area, as well as case identification numbers. Tabulated data may be available from Brant County Health Unit by request.

